# Dynamics of COVID-19 epidemics: SEIR models underestimate peak infection rates and overestimate epidemic duration

**DOI:** 10.1101/2020.04.02.20050674

**Authors:** Alastair Grant

## Abstract

Compartment models of infectious diseases, such as SEIR, are being used extensively to model the COVID-19 epidemic. Transitions between compartments are modelled either as instantaneous rates in differential equations, or as transition probabilities in discrete time difference or matrix equations. These models give accurate estimates of the position of equilibrium points, when the rate at which individuals enter each stage is equal to the rate at which they exit from it. However, they do not accurately capture the distribution of times that an individual spends in each compartment, so do not accurately capture the transient dynamics of epidemics. Here we show how matrix models can provide a straightforward route to accurately model stage durations, and thus correctly reproduce epidemic dynamics. We apply this approach to modelling the dynamics of a COVID-19 epidemic. We show that a SEIR model underestimates peak infection rates (by a factor of three using published parameter estimates based on the progress of the epidemic in Wuhan) and substantially overestimates epidemic persistence after the peak has passed.

## Introduction

Mathematical models are widely used to understand and predict the dynamics of epidemics, and to assess the likely effectiveness of different disease management measures such as quarantining of infected individuals. Standard compartment models of disease, such as SEIR, are being widely used to model the dynamics of the COVID-19 epidemic (Berger, Herkenhoff, & Mongey, 2020; Danon, Brooks-Pollock, Bailey, & Keeling, 2020; Fang, Nie, & Penny, 2020; Huang, Yang, Dai, Tian, & Chen, 2020; Kucharski et al.; Pan et al., 2020; Prem et al.; Read, Bridgen, Cummings, Ho, & Jewell, 2020; Tang et al., 2020; Teslya et al., 2020; Wu, Leung, & Leung, 2020; C. Yang & Wang, 2020; Z. Yang et al., 2020). This model groups all “exposed” and “infective” individuals into single compartments. The rate at which susceptible individuals acquire infection is a function of the number of infective individuals present (as well as infectivity and contact rates). But these models express other transitions from one compartment to the next as the reciprocal of the average time that an individual spends in the first compartment. In the simplest SEIR formulation, exposed individuals spend T_e_ days before becoming infective and infective individuals spend T_i_ days before recovering [or dying], so the transition rates from E to I and from I to R are functions of T_e_^-1^ and T_i_^-1^ respectively. So the models do not track information on the time at which an individual was exposed or when they became infective.

Under this formulation, the distribution of residence times of an individual within a stage is exponentially distributed. This is biologically unreasonable, as it means that the most probable residence time in a stage is 0 for a continuous time model and a single time step for a discrete time model. In addition, some individuals spend very much longer than the average in a model compartment than is the case in reality. These standard formulations are able to give an accurate prediction of equilibrium points, as at equilibrium the number in each compartment is time-invariant, with equal numbers entering and leaving. They are, however, unable to accurately describe the dynamics of epidemics (Krylova & Earn, 2013; Lloyd, 2001a, 2001b; Wearing, Rohani, & Keeling, 2005).

Consider, for example, the introduction of a pathogen as an impulse function to a population consisting entirely of susceptibles, as might occur in a bioterrorism event. For simplicity we assume that the resulting disease has constant incubation and infective periods. A proportion of the population will acquire the pathogen and move into the E compartment. At the end of the incubation period, these will display symptoms and become I’s. If secondary infection is not possible, not further individuals will enter into the E compartment and individuals in I will remain in this compartment for the infective period before transitioning to R, unless they die. We would like a model to preserve the shape of the initial impulse function, as it passes through the E and I compartments.

But in a continuous time formulation, some individuals will immediately move from the exposed compartment, through infected and to recovering. In a discrete time model, the same will happen over the next two time-steps. Other individuals will spend substantially longer than average in both the E and I compartments, and the pathogen will persist in the model for much longer than it will in reality. This also has important implications for disease management – the standard models with exponentially distributed stage durations over-estimate the effectiveness of quarantine as a management intervention and under-estimate the importance of early detection (Wearing et al., 2005). We provide a worked example of this in the COVID-19 modelling section below.

These problems have been recognised previously. It is possible to retain the relative simplicity of an ODE formulation by subdividing compartments into a number of sequential stages (Bailey, 1964; Lloyd, 2001b). This leads to so-called Erlang distributed models, which can be formulated using the same parameters as the ODE formulation, although the number of state variables is increased (Krylova & Earn, 2013). But extending this approach to model arbitrarily distributed stage durations requires the use of more mathematically challenging integro-differential, delay differential, or partial differential equations (Lloyd, 2001a).

## An Alternative modelling approach

An alternative approach is to use a matrix model which identifies individuals that have spent different times in the E and I compartments (in effect doing the same as Erlang distributed models), but in a way which can incorporate arbitrary distributed stage durations. Matrix models have the additional advantage that it is straightforward to incorporate stochastic process variation into the model and to model numbers of individuals in each stage as integers, which will be of particular benefit for modelling the initial stages of an epidemic. The number of individuals in each compartment, and how long they have been there, is tracked in a vector:

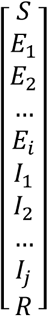

In each time step, this is multiplied by a matrix in which terms on the leading diagonal represent the probability of staying in that state (this would apply to S and R). Terms on the subdiagonal are the probabilities of moving to the next state, with the value 1 to “age” all individuals in a state by one time step (for example, to move individuals from E_1_ to E_2_ without any mortality. An expression for the probability of a susceptible acquiring infection appears in the first column of the second row with the probability of a susceptible *not* becoming infected in the top left corner. Other entries in the top row are the probabilities of moving back to being susceptible at the end of the infective period; entries in the bottom row are the probabilities of moving into the recovering compartment. If the infective period is a fixed length, then the second last element on the bottom row will be the survival rate from the last infected day to become recovering (with the bottom right hand element being the survival probability for R’s). If there is variation in the duration of the infective period, there will be non-zero entries in more of the elements on the bottom row, and variation in the duration of the latent period can be captured in a similar way. So for a model with an incubation period of 1 or 2 days, an infective period of between 3 and 4 days and no mortality, we might have (with zeros in the projection matrix and subscripts for time step in the population vector omitted for clarity):

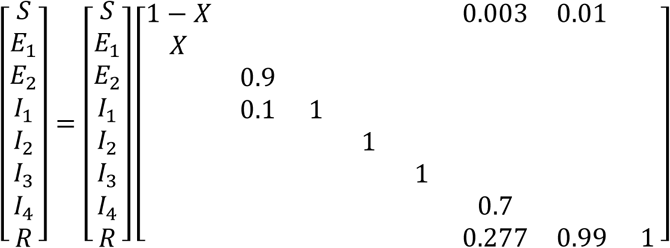

X is the probability of a susceptible being infected (which will be a function of I = ΣI_i_); 10% of exposed individuals become infective after one day, while the remainder become infective after 2 days. The infective period is 4 days in 70% of individuals, but only 3 days in the remaining 30%. There is a 1% chance of an infected individual recovering without acquiring immunity, and thus returning to the susceptible pool. Once individuals have recovered and acquired immunity, they remain in the R compartment indefinitely.

Figure 1 shows the time course of an epidemic resulting from a population consisting of 1000 of susceptibles in which a single individual has recently acquired infection. The incubation period is fixed at 4 days and the infectious period is fixed at 5 days. Infectivity is high. Each individual encounters 20 others per day, with a probability of 0.5 of acquiring the infection from each encounter with an infective. In the standard SEIR model (Fig. 1a), the number of exposed individuals has increased before day 5, even though the initial primary case is not yet infective. The number of infective individuals makes up 25% of the population by day 10, when the first secondary cases should only just have started to appear, and the number of infective individuals peaks on day 13. However, the persistence of the epidemic is relatively long, with some infective individuals still remaining beyond day 35. By contrast, the fixed stage duration model (Fig. 1b) correctly predicts that the number of exposed individuals remains low until after day 5, as secondary infection can only occur when the primary case becomes infective. There is then a linear increase in the number of exposed individuals until day 10, until secondary cases start to become infective and the infection then undergoes exponential growth. The peak number of infectives is not reached until day 19, and is higher than in the SEIR model, but the epidemic then rapidly declines, and becomes extinct on day 25.

**Figure 1a.**
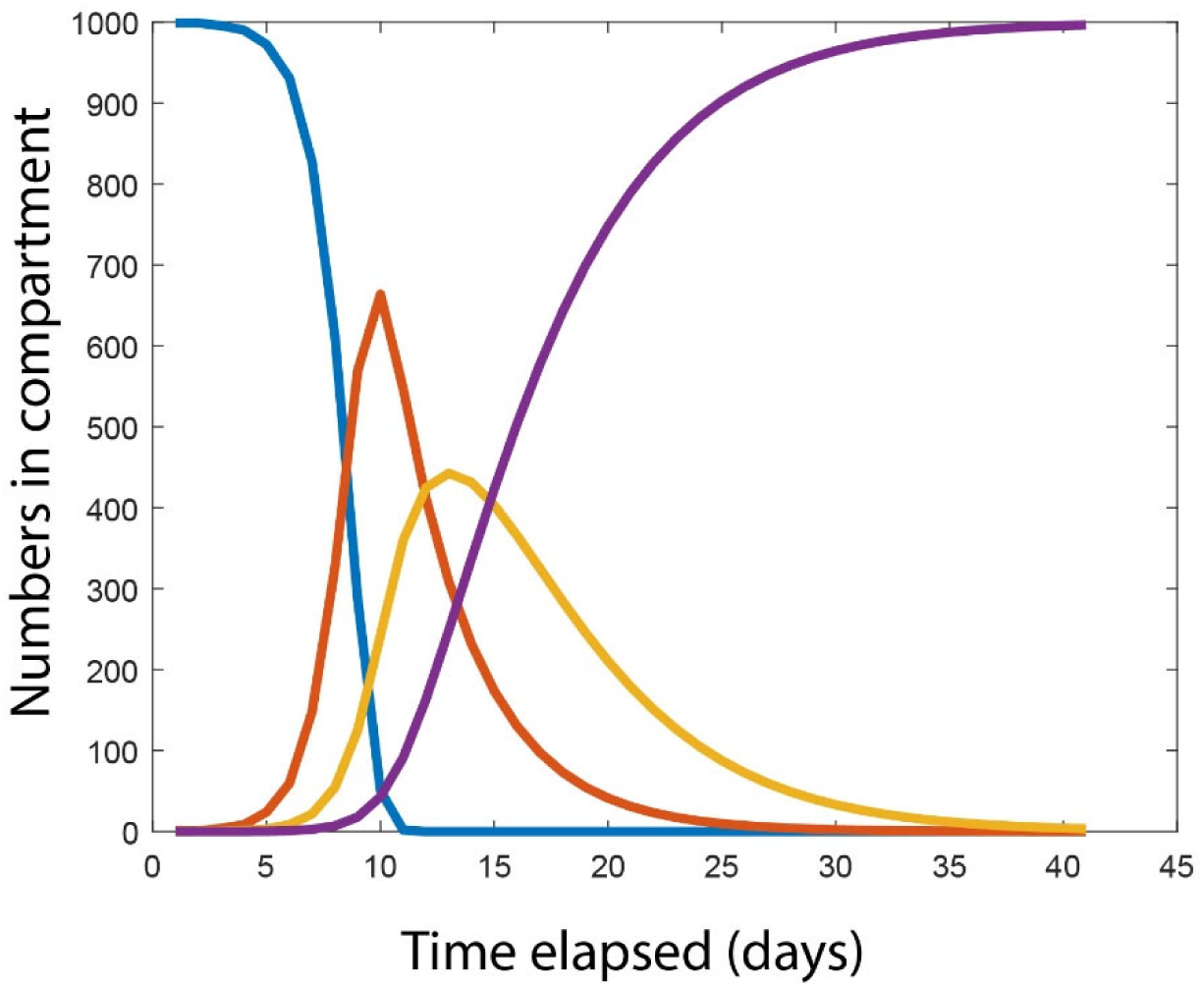
Time course of an epidemic beginning with a single exposed individual. Latent period is 4 days; infectious period is 5 days Standard discrete time. Infection probability is high (see text for details); SEIR formulation. S (Blue), E (Red), I (yellow) and R (purple)

**Figure 1b.**
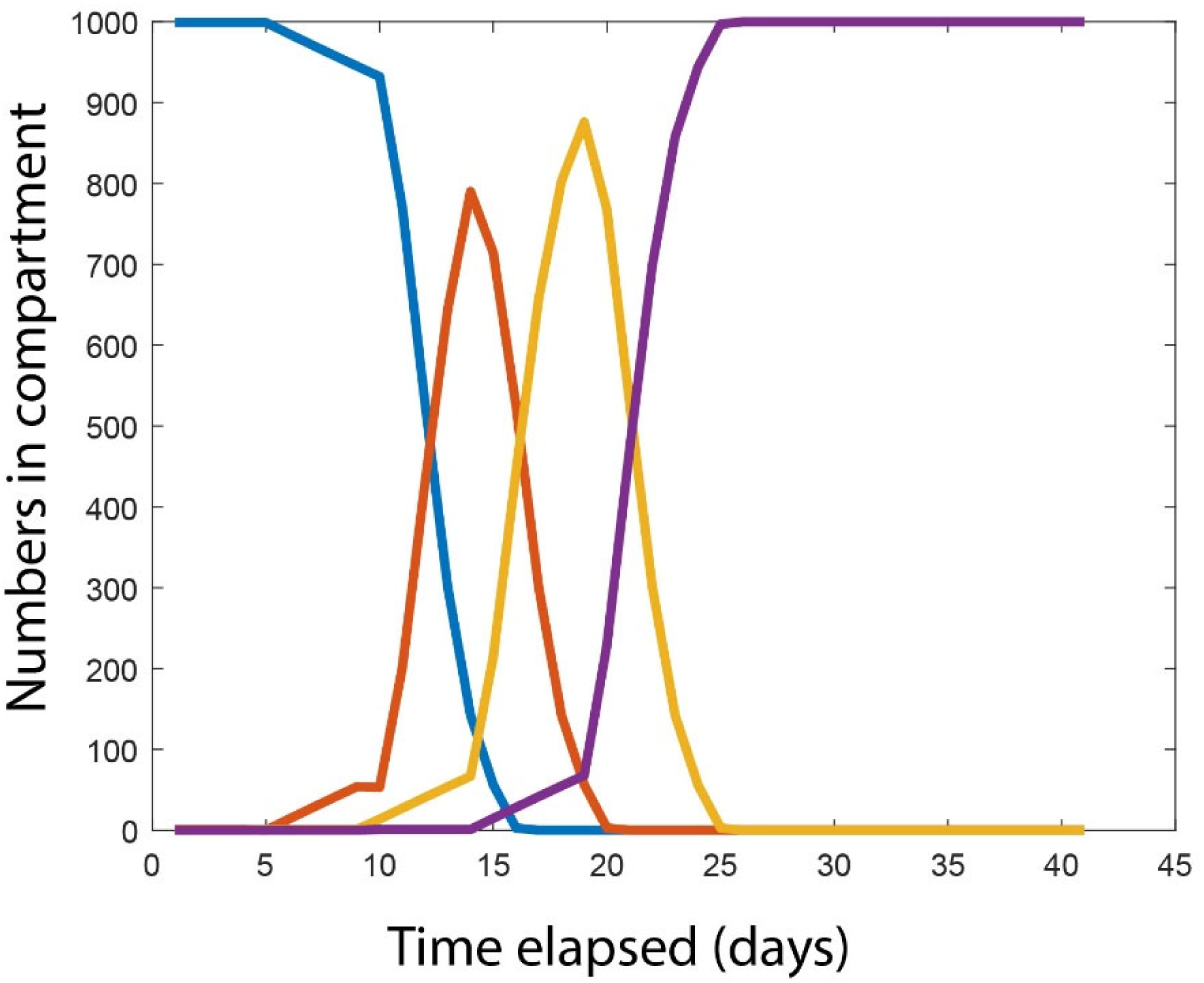
Time course of an epidemic beginning with a single exposed individual when using explicit time SEIR matrix model to maintain correct stage durations. Colour codes as Figure 1a

## An Application to COVID-19 epidemics

Next, we applying this modelling framework to a published model of the COVID-19 outbreak. Fang et al. (2020) modelled the progress of the epidemic in Wuhan City using a continuous time SEIR model:

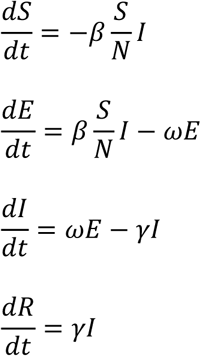

Where β, the infection rate, is given by β = β_0_k, β_0_ is the probability of infection per encounter with an infected individual and k is the number of people encountered per day. ω= 1/T_e_, where T_e_ is the latent period and γ = 1/T_i_, where T_*i*_ is the average recovery time of infectives. This formulation assumes that individuals are not infectious during the latent period. These authors used initial parameter estimates of β_0_ = 0.1; k = 10; T_e_ = 7 and T_i_ = 10.25 and a population size of 10 million. We have formulated this as a discrete time SEIR model with a time step of one day (see supplementary material). If we initiate the model with 1000 newly exposed individuals, and set secondary infection rates to zero, then the SEIR model does a rather poor job of preserving this impulse function (Fig. 2a). Infected and recovered individuals start to appear on day 2 and day 3 respectively. Numbers of infected individuals peak on day 9, but only 32% of the population is infected at this point, with 44% remaining in the E compartment and 24% already recovered. Appreciable numbers of infected individuals are still present after day 40. We have also used the same expressions for infection rates in an explicit time SEIR matrix formulation that generates a fixed length latent period (Fig. 2b), and an infective period of 10 days for 75% of individuals and 11 days for the remaining 25%. This correctly shows 100% of the population moving from exposed to infective on day 8, and all individuals recovering by day 19.

**Figure 2a.**
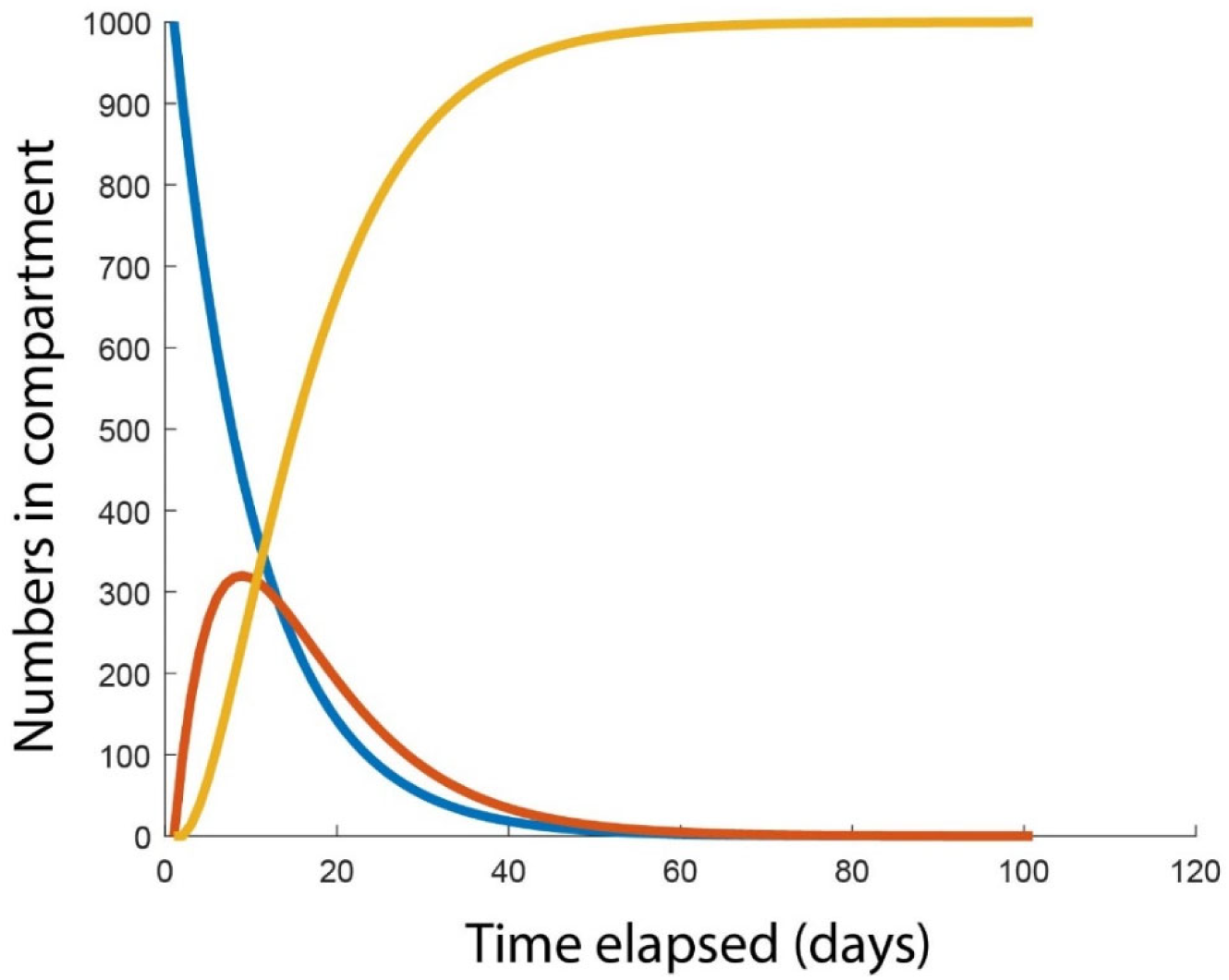
Time course of a single cohort of newly infected individuals through the E (blue), I (red) and R (yellow) compartments of a discrete time SEIR model of COVID-19 (Fang et al., 2020).

**Figure 2b.**
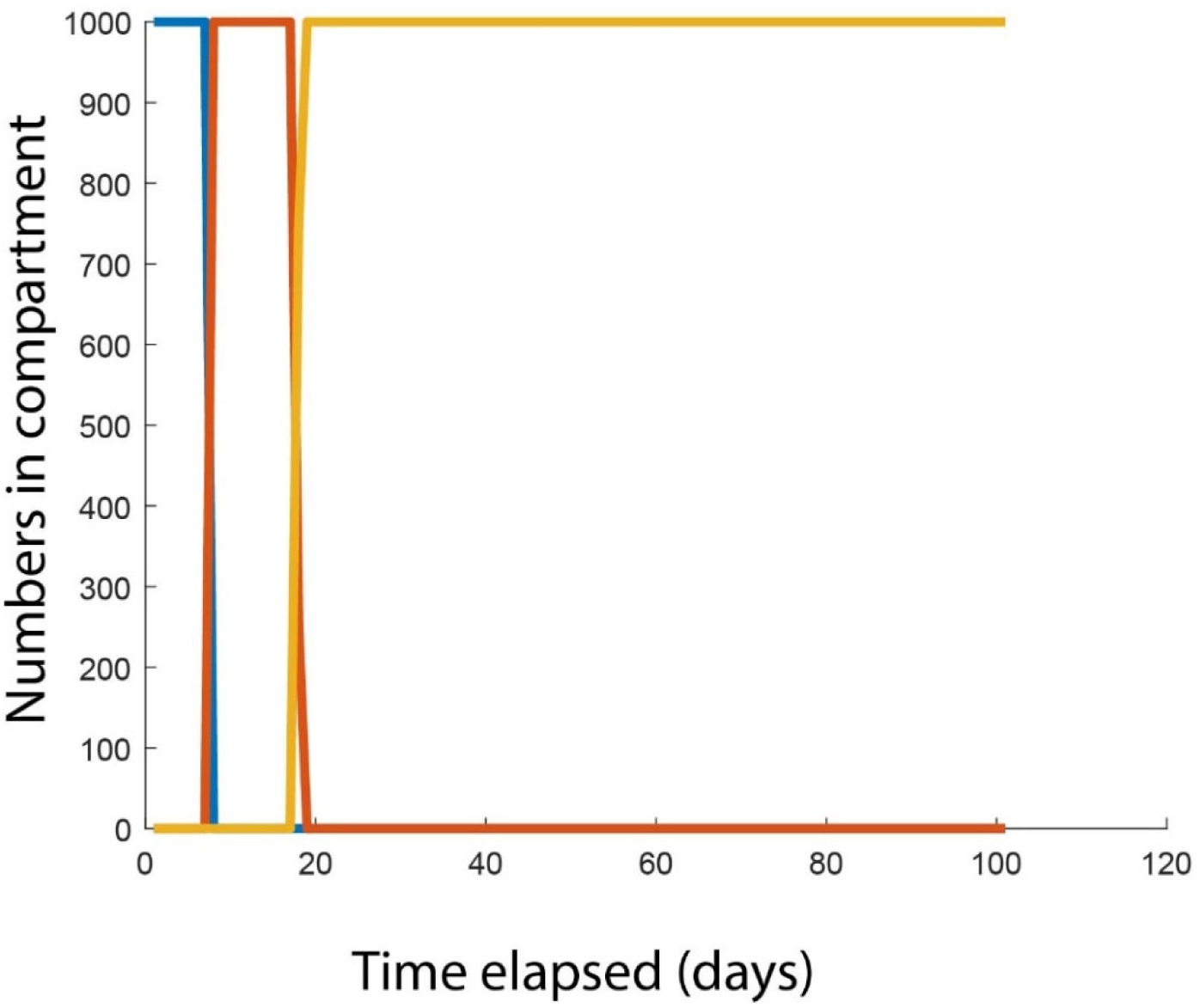
Time course of a single cohort of newly infected individuals through the E, I and R compartments of explicit time SEIR matrix model of COVID-19. Other details as in Figure 2a

So the discrete time SEIR model performs rather poorly at tracking the time course of a cohort of individuals infected at the same time. We might, therefore, expect it to perform poorly at tracking the dynamics of a rapidly growing epidemic. An epidemic that starts with a single newly exposed individual reaches a peak on day 104, with 22.8% of the population infected (Fig. 3a). The time course of the epidemic using the explicit time SEIR matrix model is shown in Figure 3b. Initial growth of the number in the I compartment is approximately twice that in the SEIR model and the peak infection rate of 63.5% of the population is reached on day 97, nearly three times the peak estimated by the SEIR model. The infection rate then rapidly declines, with only 69 infective individuals remaining by day 120 (23 days after the peak), when the SEIR model would still predict the presence of just over one million infective individuals. The number of infectives reduces to this number only by day 220 in the SEIR model (116 days after the peak).

**Figure 3a.**
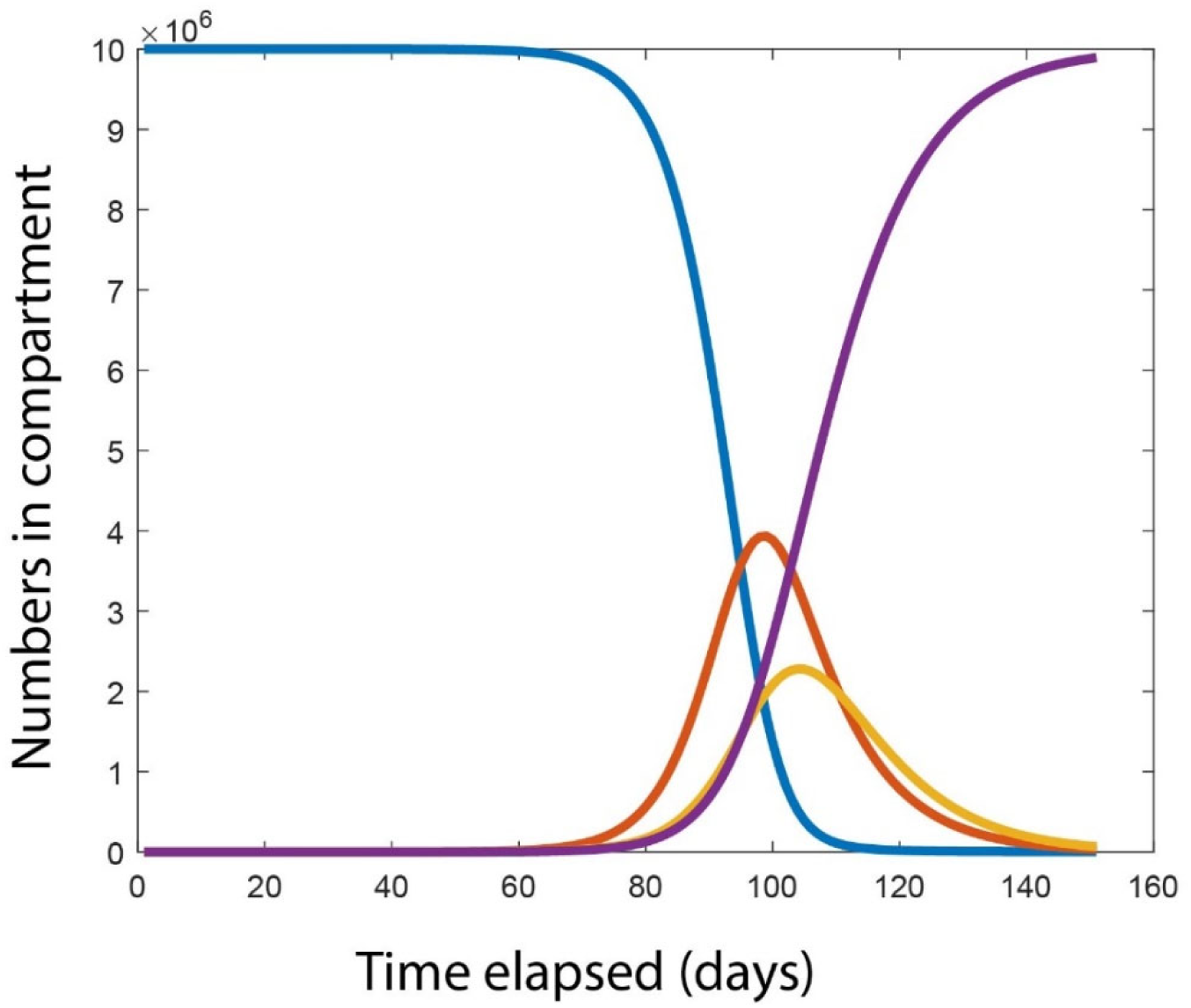
Time course of a COVID-19 epidemic beginning with a single exposed individual. Standard discrete time SEIR formulation using parameter values from (Fang et al., 2020). S (Blue), E (Red), I (yellow) and R (purple)

**Figure 3b.**
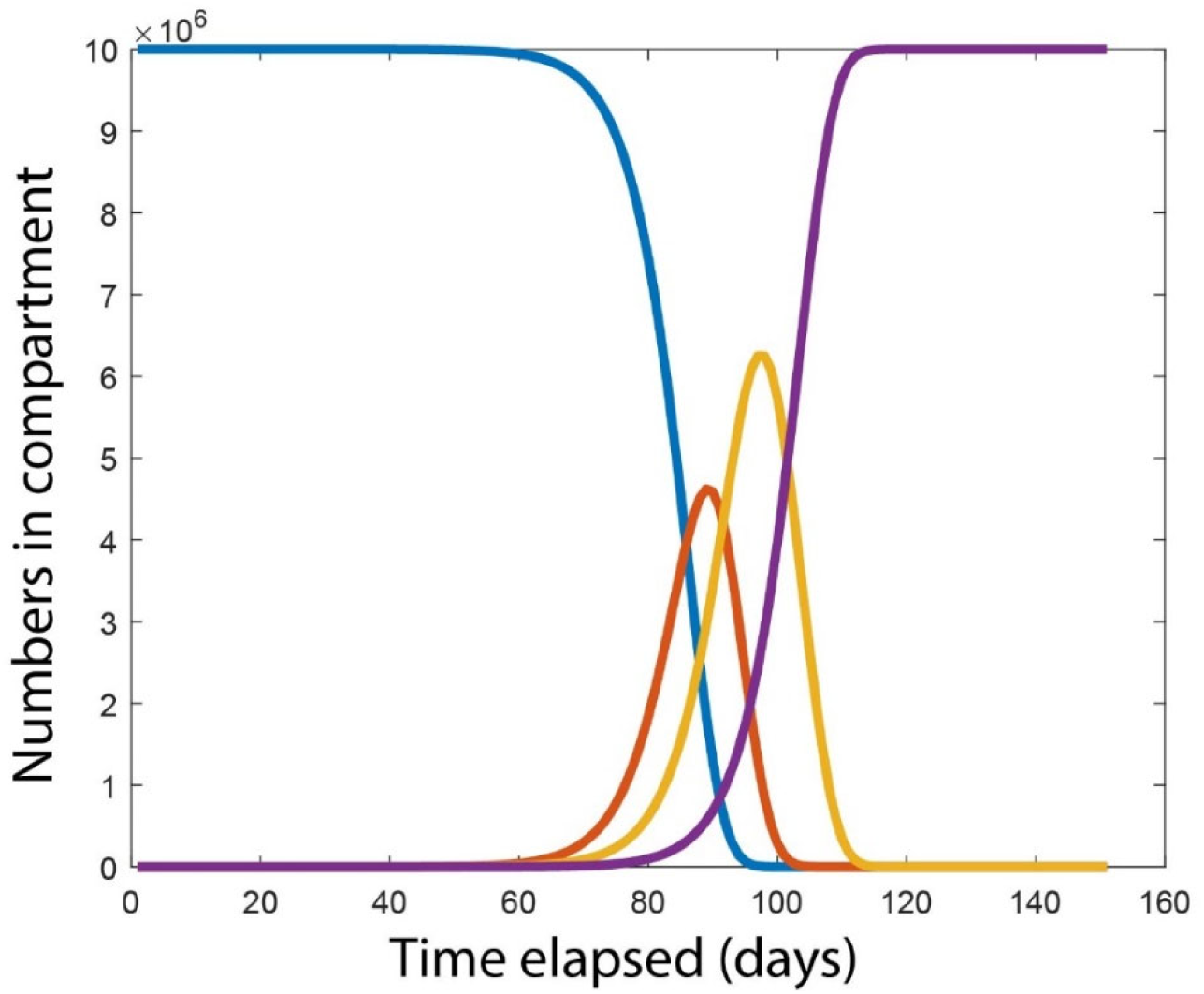
Time course of a COVID-19 epidemic beginning with a single exposed individual, modelled using explicit time SEIR matrix model. Other details as in Figure 3a.

## Discussion

We have shown that standard SEIR (and other compartment) disease models fail to correctly replicate the movement of individuals through compartments. This means that they have distributions of residence times in each stage that only resemble reality to the extent of having the same expected values, with radically different values of all higher moments. This has been known for some time (Krylova & Earn, 2013; Lloyd, 2001a, 2001b; Wearing et al., 2005) and solutions to the challenge are available (Krylova & Earn, 2013; Lloyd, 2001b). But the huge simplicity of compartment models relative to Erlang distributed; integro-differential or delay differential models has meant that they have continued to play a prominent role in disease modelling, including many published papers modelling COVID-19 (see introduction). However, their discarding of information on the length of time individuals have been in a compartment creates serious deficiencies in the ability of compartment models to replicate the dynamics of real epidemics, such as COVID-19. In the examples examined here, including a model using parameter values based on COVID-19 dynamics in Wuhan, SEIR models underestimate the proportion of the population that will be Infected when the epidemic is at its peak and overestimate the epidemic duration, by factors of three and two respectively. The models in this paper show that the peak of infection in our explicit time model may be either earlier or later than the peak in the simple SEIR model. The implications of our findings for state policies to manage of COVID-19 epidemics are not clear. If SEIR models use parameter values estimated independently from data they will underestimate the proportion of the population which will be infected at the epidemic’s peak. But if inverse modelling uses SEIR models to estimate parameters from disease time series (Fang et al., 2020), they may give parameter estimates that are too pessimistic. Another consideration is Wearing et al. (2005) assessment that the standard models overestimate the effectiveness of quarantine and underestimate the importance of early detection. It is to be hoped that national policies are being guided by a range of disease models, including ones which deal more effectively with the known time course of infection within individuals, such as agent/individual based models (Chang, Harding, Zachreson, Cliff, & Prokopenko, 2020; Ferguson et al., 2006) and time since infection models (Cori, Ferguson, Fraser, & Cauchemez, 2013; Flaxman et al., 2020; Fraser, 2007). But the domination of the published scientific literature by compartment models may be in danger of creating a discontinuity between the views held in the research community and the modelling that is informing national decision making about the management of COVID-19. We believe that the approach outlined here provides a valuable solution to the problem of incorporating a time dimension within each compartment. This allows the correct recreation of time dynamics within each compartment, while retaining the simplicity of a matrix based model and the details of the model’s assumptions can immediately be seen by examining the projection matrix. It also allows the straightforward use of tools for sensitivity/elasticity analyses of both population sizes and selection intensity on the virus characteristics (Grant, 1997; Grant & Benton, 2000). The model here could readily be adapted to incorporate infectivity during the latent period; variance in the duration of the latent and infected stages and incomplete immunity of recovered individuals, and we hope that it will be of some use to the COVID-19 modelling community.

## Data Availability

The manuscript does not refer to any data. Computer code is contained in the supplementary material

## Notes

### Competing Interest Statement

The authors have declared no competing interest.

### Funding Statement

Neither the the author or their institution have received third party funding for the work reported.

